# Influence of Illustrator’s Intentions and Visual Description Techniques of Medical Illustrations on Generating Interest and Boosting Comprehension of Medical Information

**DOI:** 10.1101/2022.04.04.22273410

**Authors:** Makiko Haragi, Ryo Onozuka, Ryosuke Nishida, Shimpei Koyama

## Abstract

In this study, an exploratory online survey was conducted to clarify points that have not been clarified so far in the medical illustration research field. If several illustrators make an illustration of the same information, does a match of intentions and techniques affect the impression or comprehension of the information? The aim is to further the utilization of medical illustration in actual practice (n=1104).

First, we selected Autosomal Dominant Polycystic Kidney Disease (ADPKD) as the medical condition about which information had to be disseminated. Then, we asked 32 art professionals to make six illustrations with three detail types (high, middle, and low) and two purposes (“getting interested” and “boosting comprehension”). Thereafter, we selected six different types of art professionals’ illustrations for the questionnaire to ask the participants about the intentions of the illustrations and comprehension of the content. We found that if the illustrator’s intentions and visual description techniques match with the recipients, the match of intentions could help generate interest in the content, and the match of visual description techniques could enhance the comprehension of the information.

## Introduction

Communication in the medical field is called health communication. In particular, a communicative relationship between doctors and patients is an essential matter (1–4). It is the prime responsibility of doctors to explain every aspect of diseases to their patients (5, 6) as excellent communication can improve the outcome of the treatment procedure (3, 4, 7–9). As most patients are not familiar with medical terminologies, occasionally, some friction emerges in the health communication field (3, 10). Previous research has tried to find efficient ways to use verbal behaviors (11, 12), attitudes (13, 14), and cultural aspects (15, 16) to provide solutions to this problem. Currently, the use of illustrations is in the limelight as a measure of effective health communication.

Illustrations used in the medical field are called medical illustrations (sometimes called infographics or visual cues) and have been used to convey medical knowledge among physicians for a long time (17). Currently, doctors also transfer detailed medical information to patients and societies. Previous research has revealed that illustrations can encourage people to remember the information (18, 19), enhance understanding (20), and secure attention (21). Moreover, illustrations are practical for someone who has low literacy, especially low medical literacy skills (18, 22, 23). Illustrations are mostly used to create concrete images of numeric information (24–26) or to depict details of diseases and affected areas comprehensively (27, 28). However, there are still plenty of things that remain to be clarified, as explained below.

In previous studies, several illustrations created by illustrators were used (29, 30). All medical illustrators cannot create the exact same illustrations as others because they would have different views, techniques, and aesthetic senses of creation (18). Considering these differences is vital for using medical illustration in the medical field to convey accurate information. However, previous studies have never conducted research using several illustrations created by various illustrators simultaneously.

Responding to the above statement, we assume that various illustrators’ illustrations would include different intentions and techniques even though the purpose of the illustrations are the same. For instance, to create a brochure to convey medical information, one medical illustrator might exaggerate some part of the information to accomplish the purpose. The result might be something abnormal or grotesque. These varying intentions and technical details have remained concealed and never had the opportunity of being revealed in previous studies. Thus, it is not clear whether these intentions and techniques could convey the information to the recipients accurately. However, we presume that if these intentions and techniques could affect the recipient’s impression and comprehension, it is worth paying attention to them with the intent of conducting more efficient health communication.

Hence, in this research, we have set up an exploratory research question as follows

RQ: When several illustrators make illustrations of the same information, does a match of intention and technique affect the impression on the recipients or their comprehension of the information?

We are convinced that studying this research question would reveal how to use medical illustration more intentionally and strategically in health communication. These perceptions would be helpful in all aspects of communication areas in the scientific field.

## Materials and methods

This study employed a quantitative design comprising an online questionnaire to evaluate the research question. Prior to the study, some art professionals created medical illustrations for different purposes. They were asked to describe their intention of illustrations and the techniques they have used. Thereafter, these responses were used for the completed questionnaire using an online survey and the research and analysis were conducted.

The survey was approved by the Ethical Review Board of Saitama Prefectural University (No. 20083), and informed consent was obtained from all participants through the online survey system. This study was conducted according to the principles of the Declaration of Helsinki.

### Selecting medical information

Before conducting the research, we selected suitable medical information. The functions of medical illustrations are affected by participant base knowledge. A previous study revealed that if recipients of the information are familiar with the medical information, the illustration would not work well to enhance comprehension compared to those who are unfamiliar recipients of the same (31). Therefore, we had to select unfamiliar medical information to reduce the distraction factors for participants.

Moreover, we focused on selecting a medical condition that is not well known generically but essential in the medical field; thus, we selected Autosomal Dominant Polycystic Kidney Disease (ADPKD), the most common hereditary kidney disorder (32, 33).

### Setting medical illustration types

After selecting the medical information to be imparted, we set the details and purposes of the illustrations. Previous studies divided illustration details into three types to understand the influences of the illustrations (29, 30). This study followed them and divided the details into three categories, as follows: realistic (high detail: HD), characteristic (medium detail: MD), and schematic (low detail: LD). Additionally, to find the “visual description techniques” of each illustrator, we divided the purposes of illustrations into two: “getting interested” and “boosting comprehension.” This point has been referenced in previous studies (27) that show that the most effective illustration, which got the recipients interested, differed from the most effective illustration, which favored comprehension. Subsequently, we made illustrators create normal drawings to clarify differences of “intentions” for each purpose. Hence, each illustrator created seven illustration types (**Figure 1**). We provided some ADPKD information, including simple exposition and pictures, to the illustrators to create the illustrations.

**Figure 1.**
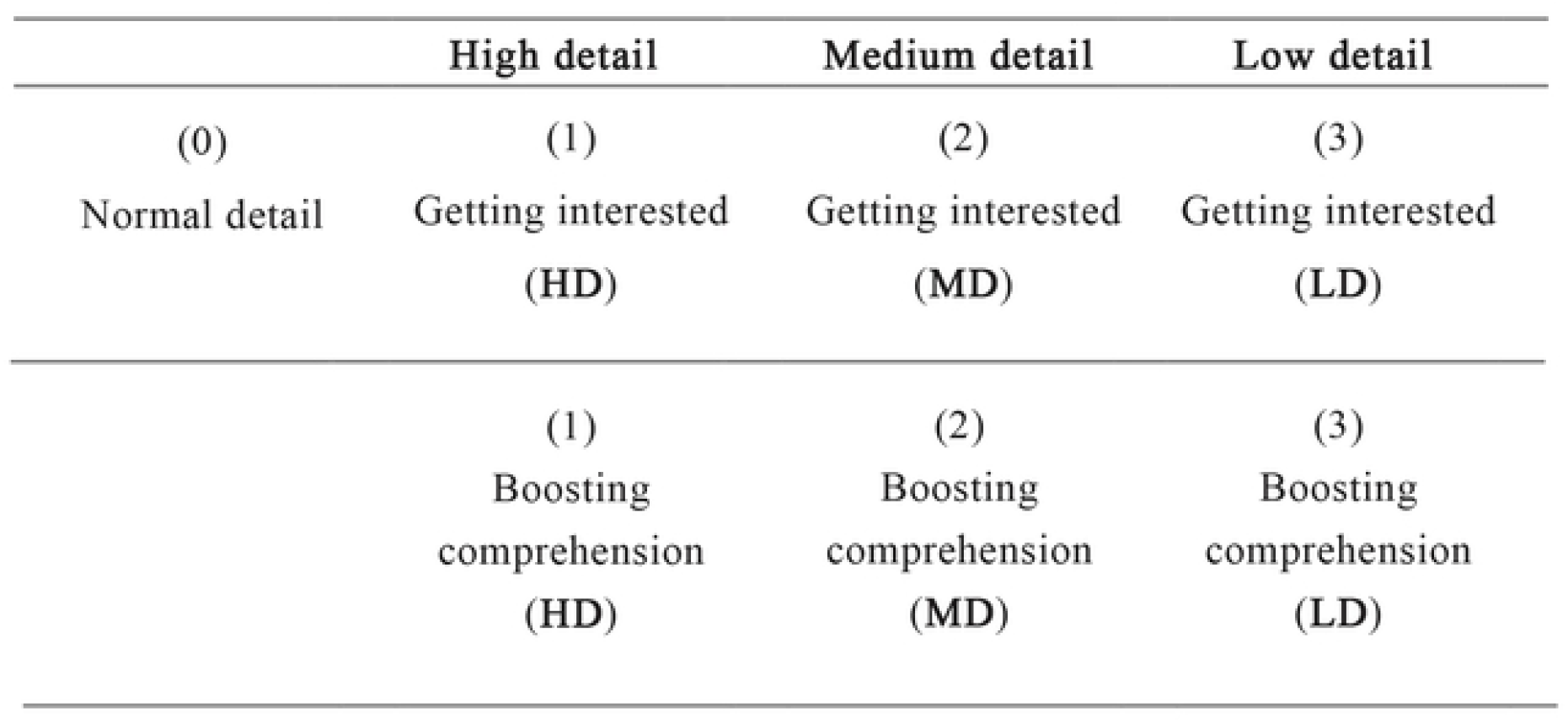
Types of illustrations drawn by an illustrator.

### Developing medical illustrations

Before the leading research, we made several medical illustrations of ADPKD. This part can be divided into two phases. First, we recruited four art professionals who have fine art college training for more than four years and have been involved in creating artworks for more than ten years in Japan. Then, we asked them to create seven types of illustration of ADPKD with two purposes, three details, and normal drawings. Afterward, we asked them to answer some questions: 1) What intentions have you included in each illustration? and 2) Point out the differences in the visual description techniques for each illustration, and provide corresponding feedback.

After modifying the methods based on the feedback and confirming the procedures, we proceeded to the second phase, gathering 35 art professionals and asking them to create ADPKD illustrations and answer questions about intentions and visual description techniques. Eventually, we collected 32 professionals and 224 illustrations.

### Picking up the illustrations’ intentions and visual description techniques

First, we selected six different types of professional illustrators for the questionnaire to find out how each illustration affects the generation of interest and comprehension of medical information related to ADPKDs (**Figure 2**). From the six illustrators’ responses to the abovementioned questions, we observed twelve keywords related to intentions (A)– and seven different types of visual description techniques (a)–(g) (**Table 1**). Then, these categories were used for the core research questionnaire to determine whether the participants of the quantitative research could detect the same intentions and visual description techniques from each illustration as professionals.

**Table 1.** Details of “Intentions” and “Visual description techniques”

| Intentions | Visual description techniques |
| --- | --- |
| (A) Painful | (a) Compare with normal and abnormal |
| (B) Feel sick | (b) Description of cross-sectional view |
| (C) Interested | (c) Description of cross-sectional |
| (D) Bizarre | (d) Description of progression of disease |
| (E) Grotesque | (e) Exclude lurid |
| (F) Anguished | (f) Description of simplicity |
| (G) Abnormal | (g) Description of features |
| (H) Surprised |  |
| (I) Fear |  |
| (J) Vitality |  |
| (K) Creeps |  |
| (L) A sense of danger |  |

**Figure 2.**
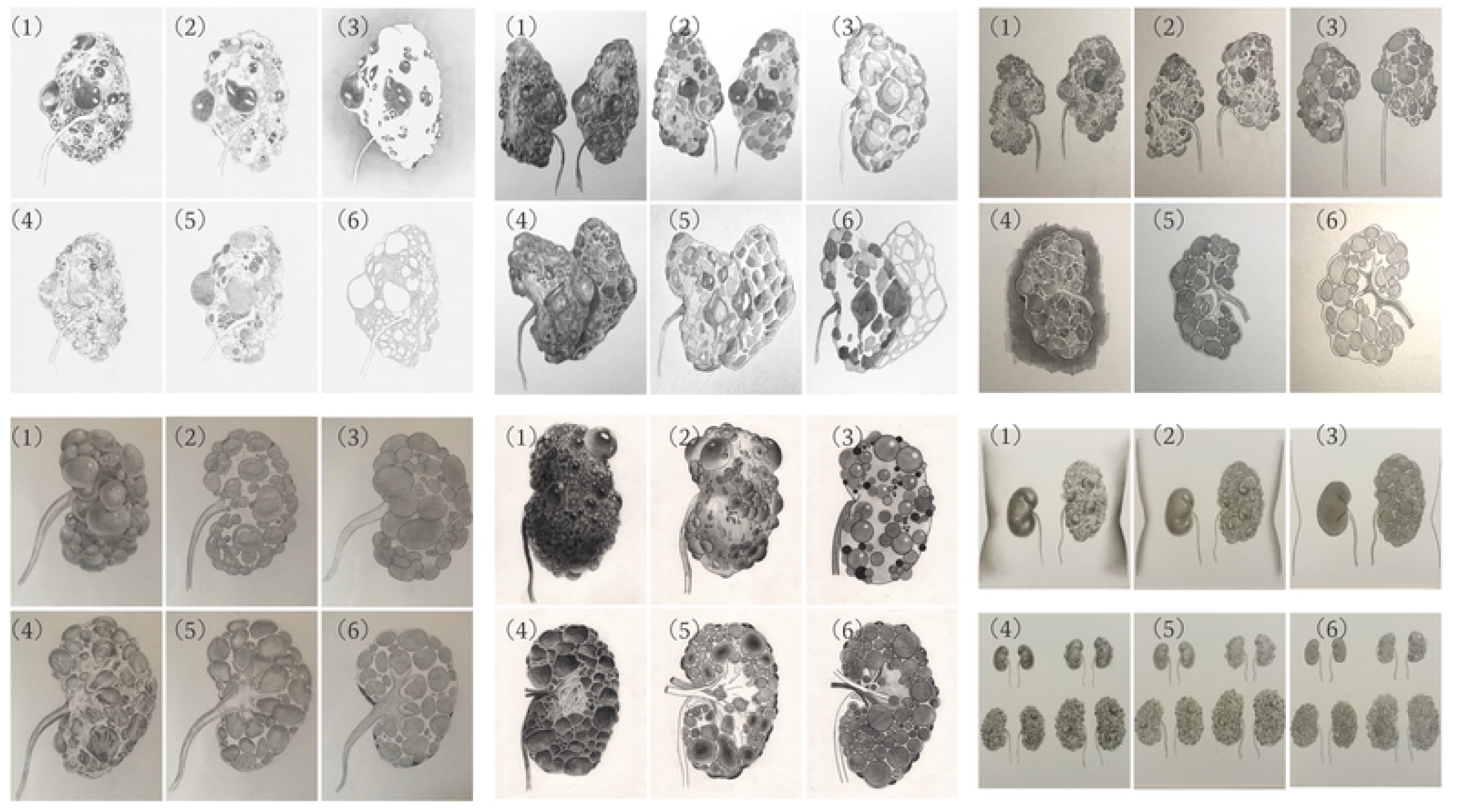
Illustrations created by six art professionals.

In the questionnaire, we showed illustrations created with three details and purpose by each illustrator. We asked the participants which illustration was the most effective for getting attention/boosting comprehension and made them pick the correct one. Afterward, we asked the participants about the visual description techniques and intentions for picking the illustrations by using (a)–(g) and (A)–(M), which were created from the art professionals’ answers. In this part, participants could choose applicable items without limitation. In addition to the above questions, the participants answered some preliminary questions on medical history and demographics before completing the questionnaire.

### Contents of the questionnaire

We used the following questions to gather information on the illustrations that generated the most interest and those that were the most comprehensible, and the participants’ impression of the intentions and visual description techniques.

(**Q1**) Which illustration did you feel succeeded in getting you interested the most when accompanied by text? (**Q2**) Which illustration was the most comprehensible when accompanied by text? Participants chose each suitable one from the three types of illustrations (realistic, characteristic, and schematic) with respect to each illustrator’s creations.

Question 1’s dependent variable was operationalized as a 7-point Likert scale from “not at all interested” to “very interested,” and the dependent variable of question 2 used a 7-point Likert scale ranging from “did not boost comprehension at all” to “deepened comprehension very much.”

In addition, with regard to each selected illustration, participants were asked to select the intentions and techniques from (A)–(L) and (a)–(g). In this part, participants could choose applicable items without limitation.

### Procedure and measures

In this research, we selected Macromill, the largest online survey service in Japan, to conduct the survey. The survey was conducted by randomly selecting a sample of 1000 respondents among the registered respondents on Macromill (500 male and 500 female). For each gender, 100 participants were assigned for each of the age categories: 20s, 30s, 40s, 50s, and over 60s. All participants were screened based on the condition that none of them are health care workers and not involved in the medical care field. We also asked demographic questions about marriage, children, household income, personal income, and occupation.

We then asked about confounding factors that might affect their interest in and understanding of the illustration. These included the respondent’s and family’s experience of hospitalization, medical history, kidney function and shape, knowledge of the disease, methods of obtaining medical information, media preference, and work experience in illustration.

After the above questions, participants were presented with some information of ADPKD, including simple expositions and pictures.

### Analysis

First, regarding the degree of correspondence between the illustrator’s intentions and the respondents’ comprehension, which is the independent variable, the following is to be noted.

This study has twelve intention categories, and an illustration can be mapped as a point in a twelve-dimensional space. For example, when an illustrator tries to convey “(A) Painful” and “(B) Feel sick,” the intended intention can be expressed as vector a in Equation 1. When the respondent answers “(A) Painful” or “(C) Interested,” the intention received can be expressed as vector b in Equation 2. The cosine of the angle between these two vectors is called the cosine similarity. If we use Equation 3, we can obtain it as a numerical value in an interval of −1, where the degree of correspondence is the least consistent, to 1, where the degree of correspondence between the intended impression and received impression is the most consistent.

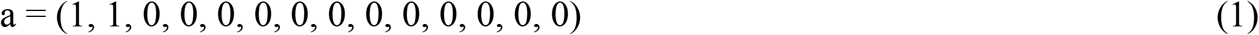

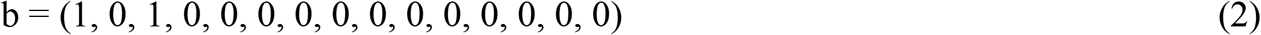

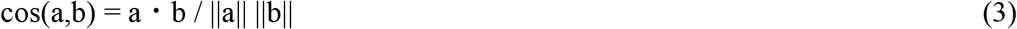

In the above sample, the cosine similarity is 0.5, and the angle between vector a and vector b is 60 degrees, which means they are relatively similar. The same formula was applied to the items in the right column of Table 1 for the degree of correspondence in the description.

Next, in addition to the single numerical value of the agreement, a dummy variable was established to specifically observe which items related to intentions and visual description techniques were in agreement. For example, the variable is 1 if the respondent received the intention of “Painful” for an illustration that was intended to convey the intention of “Painful,” and 0 otherwise. The third independent variable was the one that asked the participants to choose which of the three types of illustration (HD, MD, or LD) was the best.

Using this analysis method, we approximated the dependent variables as continuous values (Norman, 2010), and statistical analysis was conducted using normal linear models, and four models were constructed. The first model used response data about getting interested, with the degree of correspondence between intentions and visual description techniques as the independent variables.

The second model used response data about the comprehension of information, with the degree of correspondence between intentions and visual description techniques as the independent variables.

The third model used response data about getting interested and used the dummy variable for the correspondence between intentions and visual description techniques as the independent variables.

The fourth model used responses on comprehension of information, with dummy variables for intentions and visual description techniques as independent variables. All models contained confounding variables, in addition to the previously mentioned items, the type of details (HD, MD, or LD) as well as illustrators. The coefficients of each variable in the above models were interpreted at the 5% level of significance.

## Results

### Sample descriptive statistics

The total number of participants was N = 1104 (male = 555, female = 549). Other demographic items are shown in **Table 2**.

**Table 2.**
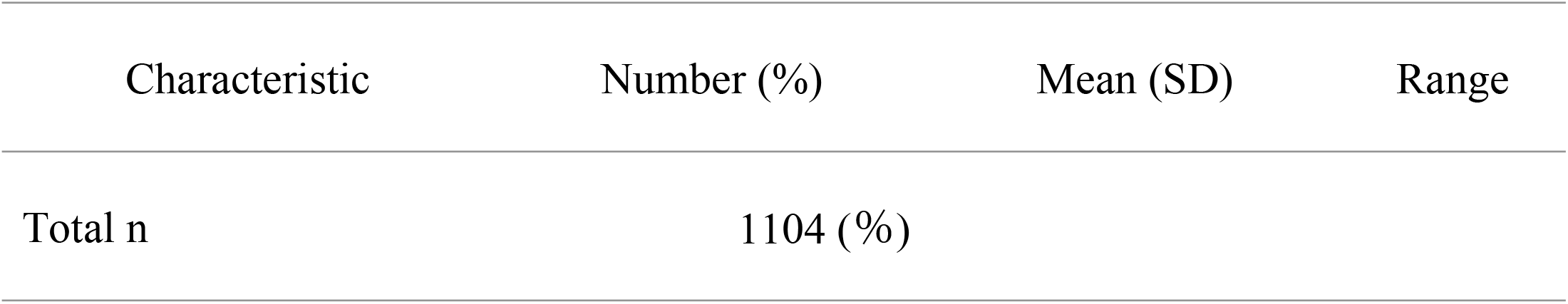
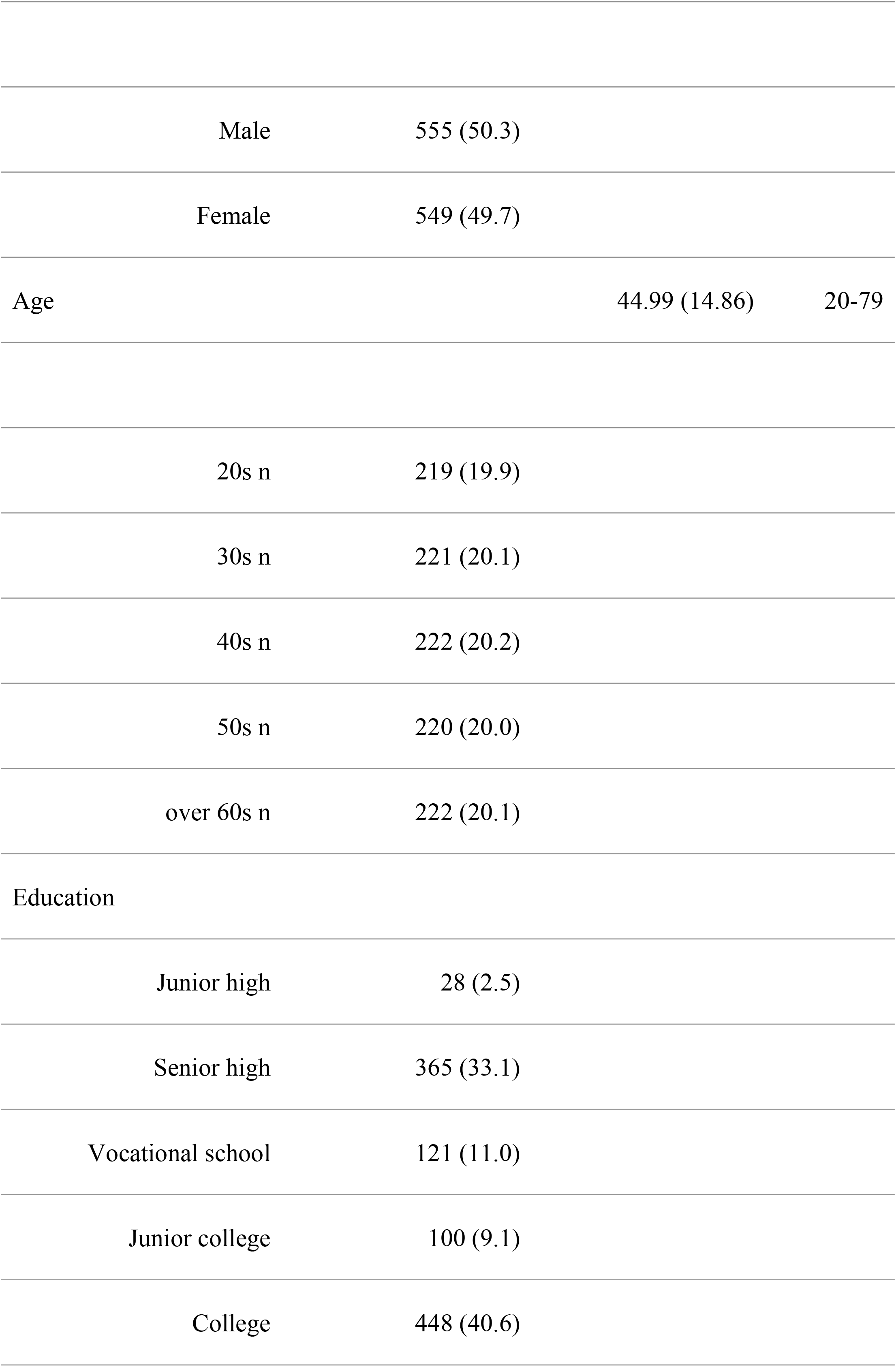
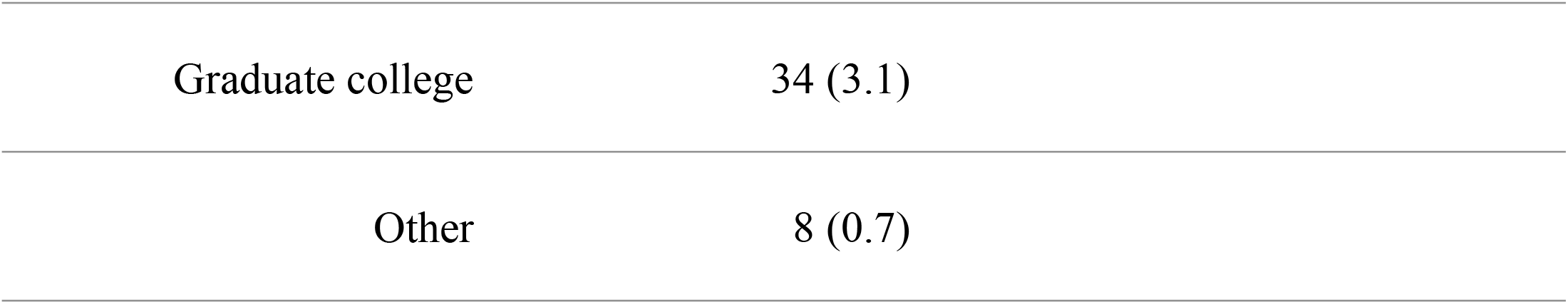
Participant details

### Summary of the models in the research

The following is a summary of the four statistical models created by this study (**Table 3**). The dependent variable is the degree of interest; in Models 1 and 2, the independent variables are the degree of correspondence of intentions and the degree of correspondence of visual description techniques; in Models 3 and 4, they are dummy variables for matched intentions and visual description techniques between illustrators and respondents; Models 1 and 3 use data about getting interested, and Models 2 and 4 use data about promoting comprehension. The confounding variables were included in the models only to ensure the reliability of the coefficients of the explanatory variables. Hence, their description is omitted in Table 3.

**Table 3.**
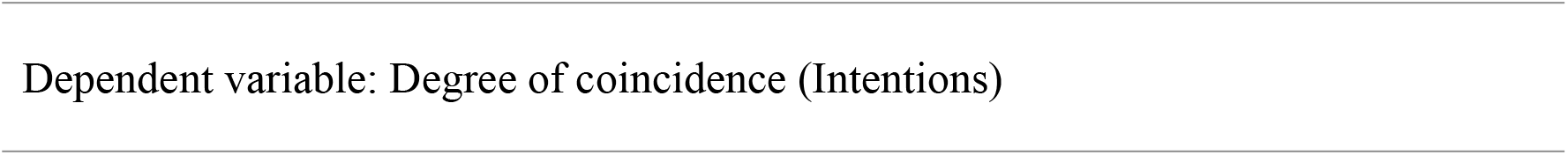
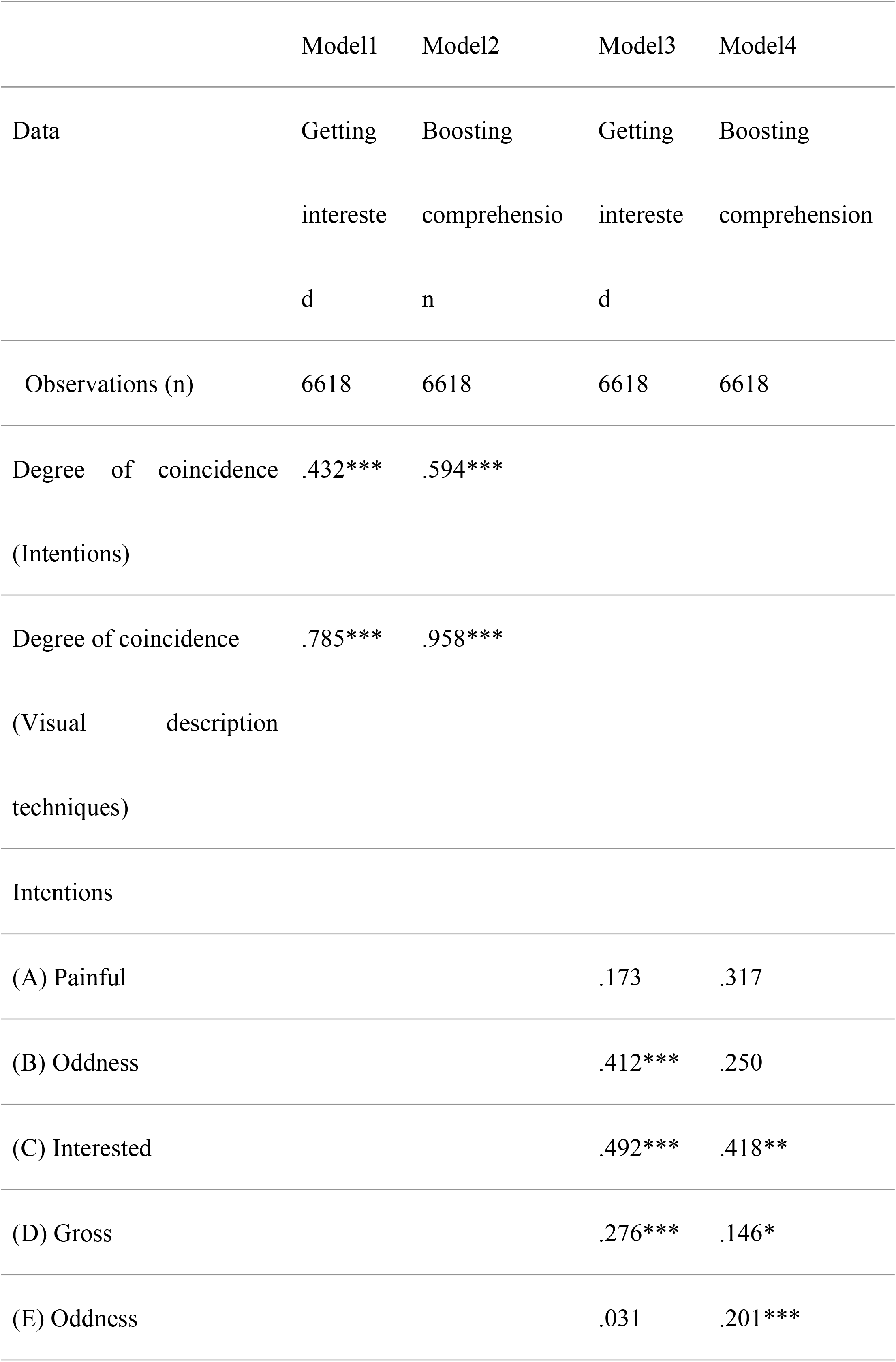
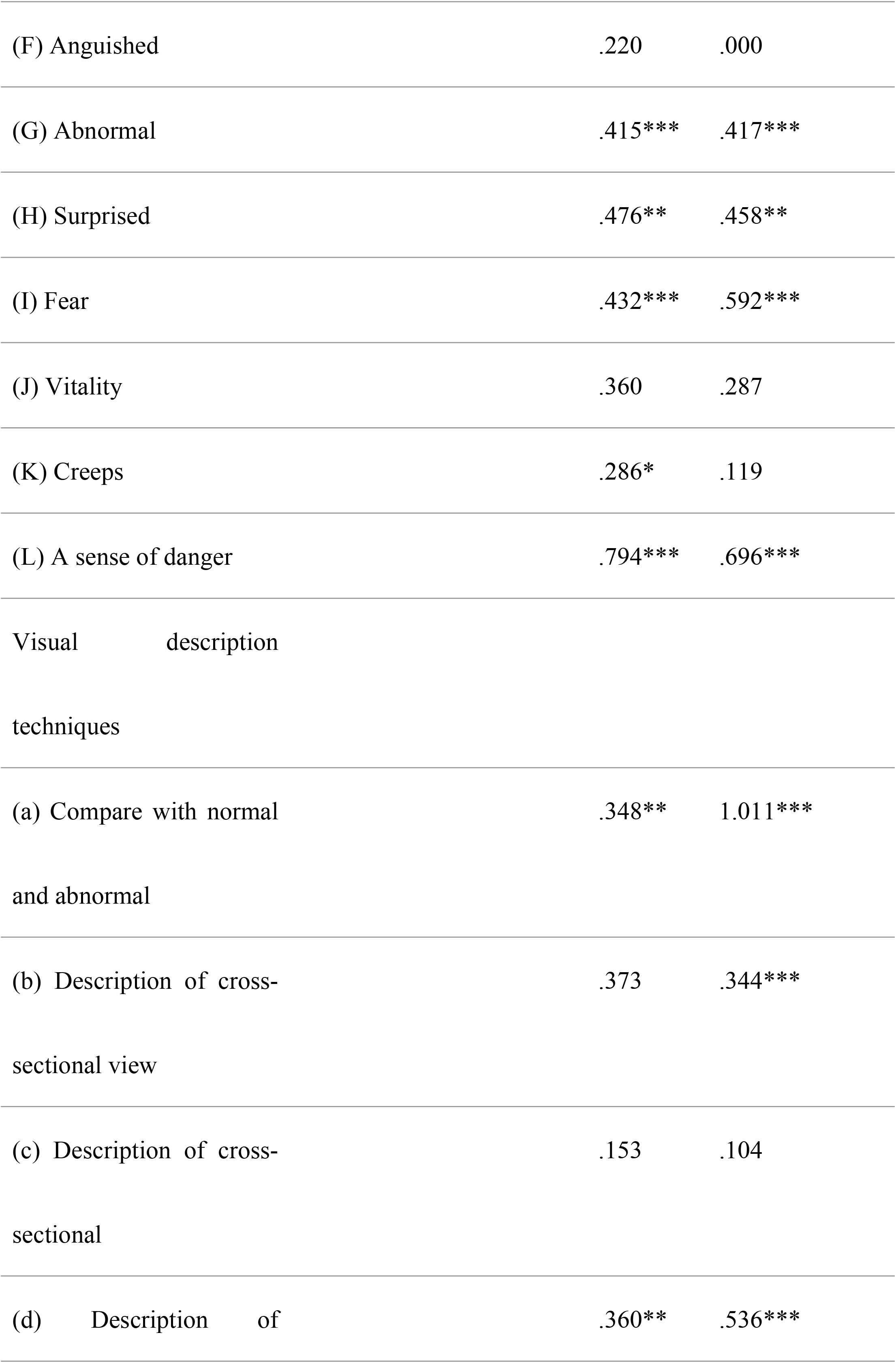
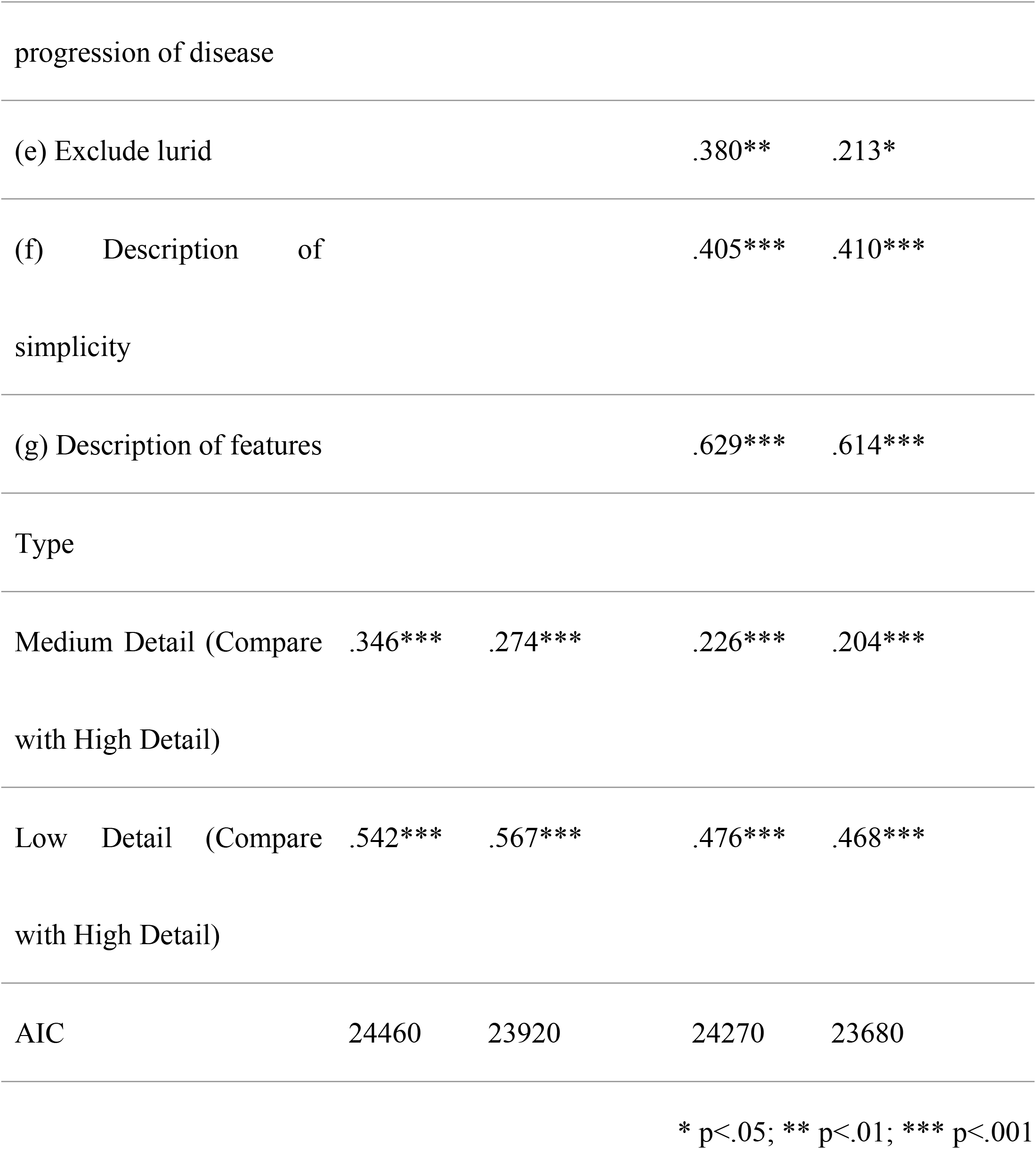
Summary of the model

### Result of Models 1–4

According to the table’s result, we focus on Models 1 and 2 first. For both models—getting interested and promoting comprehension—the degree of correspondence between illustrators and participants’ intentions and visual description techniques had a significant effect. Additionally, the degree of descriptive correspondence had a stronger effect. Moreover, both coefficients were greater than 0.1 for promoting comprehension rather than getting interested.

Next, regarding Model 3, in terms of getting interested, the intentions that had a significant positive impact on the level of interest were, in descending order of coefficient, Intentions (L), (C), (H), (I), (G), and (D). In contrast, the intention that had a negative impact was Intention (B). In terms of visual description techniques, (g), (f), (e), (d), and had a positive influence, in that order.

In Model 4, the intentions that had a significant positive impact on the degree of interest in promoting comprehension were, in order of increasing coefficient, Intentions (L), (I), (H), (C), (G), (E), and (D).

As for visual description techniques, (a), (g), (f), (d), (b), and (e) had a significant influence, in that order.

From these, the distinctive results for getting interested are the magnitude of the negative impact of Intention (B), the magnitude of the coefficient of Intention (D), and the magnitude of the coefficient of visual description techniques (e). In contrast, the features that stand out for promoting comprehension are the large coefficients of Intentions (E) and (I), the large difference in the coefficient of visual description techniques *a*, and the large coefficient of (d).

### Result differences between illustration details and interests

We discuss the coefficients against the degree of interest for the three types of illustrations (HD, MD, and LD). As these three were treated as categorical variables, they are calculated as the coefficients of the other depictions compared to the HD. The results showed that the HD had the most positive impact on the level of interest. The MD had a lower impact, and the LD had the lowest impact.

## Discussion

### The key findings of this study are described below

First, with regard to the relationship between the illustrator’s intentions and the respondent’s reception, the finding was that interest was more enhanced when the techniques used in the illustration were correctly conveyed than when what was intended to be expressed in the illustration was conveyed as intended. When comparing getting interested and promoting comprehension, correspondence of the visual description techniques was more critical in the latter In this research, we found that interest decreased significantly when negative intention (i.e., “(B) Feel sick”) was conveyed to the respondents. In contrast, we found that to promote comprehension of disease-related information by using illustrations, it is important to correctly convey the method of visual description techniques; in particular, “(a) Compare with normal and abnormal” was found to be remarkably important. Additionally, there were six intentions (C, D, G, H, I, L) and four visual description techniques (d, e, f, g) as common positive factors for both getting interest and promoting comprehension. Moreover, the HD had the most positive impact on the level of interest. The MD had a lower impact, and the schematic depiction had the lowest impact in this study.

### Effect of intention matches

First, we would like to discuss the effect of negative intentions. This research showed that negative intentions (i.e., “(B) Feel sicky”) decrease the interest in the content. This result could be connected with a previous study’s viewpoint (34) that an uncomfortable feeling from seeing visual material could generate disinterest in the content.

As the goal of this study is to accurately communicate medical information to patients and laypersons and maintain interest in the information, we could say that the use of uncomfortable visual descriptions is not desirable. Unfortunately, this research could not reveal where the impression of “(B) Feel sick” stemmed from. It might come from visual descriptions of the illustration or it may depend on the participant’s experiences and characteristics. Therefore, it could be said that further research is needed to explain these points in more detail.

Meanwhile, another negative intention, “(L) A sense of danger,” is one of the most important factors to getting interested and promoting comprehension of content. This result may be possibly related to the function of graphic warning labels such as those on cigarette packs. Previous researches showed that when people get aware of some risks, they try to avoid potentially dangerous activities (35, 36). Therefore, there is some possibility that the impression of “(L) A sense of danger” could be used to encourage people to get interested and promote comprehension of the content to avoid some critical situations.

Applying these two points for the RQ of this study, we could say that if the illustrator’s intentions match the recipients, especially if the negative intentions match, then there is a possibility of affecting content interest. Notably, an awkward intention—” (B) Feel sick”—would have the potential influence of decreasing interest in the content, whereas intentions related to the individual health situation, like “(L) A sense of danger and (I) Fear,” could be effective for not only generating interest but also boosting comprehension of the content.

Therefore, it can be suggested that conveying a sense of risk to an individual might be useful to boost interest in and understanding of medical information content. However, it is still difficult to generalize these points, and we need to determine how to achieve these results in medical illustrations using different medical information. To standardize these results, further research is required.

### Effect of visual description technique matches

We would also like to mention the impact of the match of visual description techniques. In this study, we used illustrations created by six art professionals, which were composed of a composite of seven visual description techniques. Among them, when illustrators’ and recipients’ cognition matched using “(a) Compare with normal and abnormal,” comparing normal kidneys and ADPKD-affected kidneys had a strong impact on promoting comprehension of content. Additionally, matching using “(d) Description of progression of disease” was also found to be effective in promoting comprehension. Moreover, in “(g) Description of features,” the characteristics of the kidney ware emphasized, which, in terms of illustration detail, was found to be effective in both getting interest and promoting comprehension of its content.

This study is the first one to divide the visual descriptions technique into several factors and investigate whether a match of visual description techniques with recipients can affect the impression or comprehension of information. Therefore, these findings have possibly become new perceptions in this research field.

We also assumed that we could only mention some techniques related to the types of illustration detail using previous research results. Some previous studies showed that HD illustrations are the most efficient to boost comprehension of the content. Hence, using a technique of “(g) Description of features,” it could be possible to regard the use of HD for illustration as the most suitable method of conveying medical information to patients and laypersons.

Taking together these results, we could say that if the illustrator’s visual description techniques match recipient perceptions, techniques about disease transformation, like “(a) Compare with normal and abnormal” and “(d) Description of progression of disease,” could be effective to boost comprehension of the information.

### Limitations

There are some limitations to this research.

First, this research has an exploratory design and was conducted using certain medical information. Therefore, the results could not be standardized and need further research using various medical information for confirmation.

Second, we recruited some art professionals to create medical illustrations, most of whom have no training as medical illustrators. Therefore, whether using those illustrations for research was desirable or not, is a controversial issue.

Third, this research gathered participants online. It cannot be confirmed whether this participants’ group represents our society correctly. With a change in groups, the results have the possibility of changing.

## Conclusion

In this study, we focused on the function of medical illustrations to convey medical information to contribute smooth health communication between doctors or researchers and patients or laypersons and set an exploratory RQ that includes new points of view about illustrations’ intentions and visual description techniques.

As a result, we found that if the illustrator’s intentions and visual description techniques match with the recipients, this could help generate interest in the content, and a match of visual description techniques could enhance the comprehension of the information. These results have not been standardized yet and only apply to a particular kind of medical information and specific population group. Still, it can be said that it would be possible to use these results for actual practices experimentally and for further exploratory research.

## Data Availability

Data cannot be shared publicly because of including Illustrations drawn using analog techniques. Data are available from the Ethics Committee (contact via) for researchers who meet the criteria for access to confidential data.

## Acknowledgments

The authors thank all participants, art professionals, and the technical support team at Macromill Inc.

## References

1. Petracci M, Schwarz PK, Sanchez Antelo VIM, Mendes Diz AM. Doctor–patient relationships amid changes in contemporary society: A view from the health communication field. Health Sociology Review. 2017;26(3):266–79.

2. Kurtz SM. Doctor-patient communication: principles and practices. Canadian Journal of Neurological Sciences. 2002;29(S2):S23–S9.

3. Ong LM, De Haes JC, Hoos AM, Lammes FB. Doctor-patient communication: a review of the literature. Social science & medicine. 1995;40(7):903–18.

4. Ha JF, Longnecker N. Doctor-patient communication: a review. Ochsner Journal. 2010;10(1):38–43.

5. Brown RS, Peikes D, Peterson G, Schore J, Razafindrakoto CM. Six features of Medicare coordinated care demonstration programs that cut hospital admissions of high-risk patients. Health affairs. 2012;31(6):1156–66.

6. Timmermans S. The engaged patient: The relevance of patient–physician communication for twenty-first-century health. Journal of Health and Social Behavior. 2020;61(3):259–73.

7. Riedl D, Schüßler G. The influence of doctor-patient communication on health outcomes: a systematic review. Zeitschrift für Psychosomatische Medizin und Psychotherapie. 2017;63(2):131–50.

8. Ward P. Trust and communication in a doctor-patient relationship: a literature review. Arch Med. 2018;3(3):36.

9. Jahan F, Siddiqui H. Good Communication between Doctor-Patient Improves Health Outcome. European Journal of Medical and Health Sciences. 2019;1.

10. Thompson CL, Pledger LM. Doctor-patient communication: is patient knowledge of medical terminology improving? Health Communication. 1993;5(2):89–97.

11. Rowland-Morin PA, Carroll JG. Verbal communication skills and patient satisfaction: A study of doctor-patient interviews. Evaluation & the health professions. 1990;13(2):168–85.

12. Vogel D, Meyer M, Harendza S. Verbal and non-verbal communication skills including empathy during history taking of undergraduate medical students. BMC medical education. 2018;18(1):1–7.

13. Matusitz J, Spear J. Effective doctor–patient communication: an updated examination. Social work in public health. 2014;29(3):252–66.

14. Marcinowicz L, Konstantynowicz J, Godlewski C. Patients’ perceptions of GP non-verbal communication: a qualitative study. British Journal of General Practice. 2010;60(571):83–7.

15. Schouten BC, Meeuwesen L. Cultural differences in medical communication: a review of the literature. Patient education and counseling. 2006;64(1-3):21–34.

16. Schouten BC, Meeuwesen L, Tromp F, Harmsen HA. Cultural diversity in patient participation: the influence of patients’ characteristics and doctors’ communicative behaviour. Patient education and counseling. 2007;67(1-2):214–23.

17. Hajar R. Medical illustration: art in medical education. Heart views: the official journal of the Gulf Heart Association. 2011;12(2):83.

18. Houts PS, Doak CC, Doak LG, Loscalzo MJ. The role of pictures in improving health communication: a review of research on attention, comprehension, recall, and adherence. Patient Educ Couns. 2006;61(2):173–90.

19. Garcia-Retamero R, Okan Y, Cokely ET. Using visual aids to improve communication of risks about health: a review. The Scientific World Journal. 2012;2012.

20. Brotherstone H, Miles A, Robb KA, Atkin W, Wardle J. The impact of illustrations on public understanding of the aim of cancer screening. Patient Educ Couns. 2006;63(3):328–35.

21. Delp C, Jones J. Communicating information to patients: the use of cartoon illustrations to improve comprehension of instructions. Acad Emerg Med. 1996;3(3):264–70.

22. Choi J. Literature review: using pictographs in discharge instructions for older adults with low‐literacy skills. Journal of clinical nursing. 2011;20(21‐22):2984–96.

23. Park J, Zuniga J. Effectiveness of using picture-based health education for people with low health literacy: An integrative review. Cogent Medicine. 2016;3(1):1264679.

24. Apter AJ, Paasche-Orlow MK, Remillard JT, Bennett IM, Ben-Joseph EP, Batista RM, et al. Numeracy and communication with patients: they are counting on us. J Gen Intern Med. 2008;23(12):2117–24.

25. Garcia-Retamero R, Hoffrage U. Visual representation of statistical information improves diagnostic inferences in doctors and their patients. Social Science & Medicine. 2013;83:27–33.

26. Garcia-Retamero R, Cokely ET, Hoffrage U. Visual aids improve diagnostic inferences and metacognitive judgment calibration. Frontiers in Psychology. 2015;6:932.

27. Haragi M, Hayakawa M, Watanabe O, Takayama T. An exploratory study of the efficacy of medical illustration detail for delivering cancer information. Journal of Visual Communication in Medicine. 2021;44(1):2–11.

28. Haragi M, Yamaguchi R, Okuhara T, Kiuchi T. Questionnaire survey of a mock jury on their impressions of medical-legal illustrations aimed at reducing trauma and PTSD of jurors. Journal of Visual Communication in Medicine. 2020;43(2):67–75.

29. Strong J, Erolin C. Preference for detail in medical illustrations amongst professionals and laypersons. J Vis Commun Med. 2013;36(1-2):38–43.

30. Haragi M, Ishikawa H, Kiuchi T. Investigation of suitable illustrations in medical care. J Vis Commun Med. 2019:1–11.

31. Levin JR. On functions of pictures in prose. In: Pirozzolo FJ, and Wittrock, M. C., editor. Neuropsychological and Cognitive Processes in Reading. New York: Academic Press; 1981. p. 203–28.

32. Lanktree MB, Haghighi A, Guiard E, Iliuta I-A, Song X, Harris PC, et al. Prevalence estimates of polycystic kidney and liver disease by population sequencing. Journal of the American Society of Nephrology. 2018;29(10):2593–600.

33. Willey CJ, Blais JD, Hall AK, Krasa HB, Makin AJ, Czerwiec FS. Prevalence of autosomal dominant polycystic kidney disease in the European Union. Nephrology Dialysis Transplantation. 2017;32(8):1356–63.

34. Isen AM, Noonberg A. The Effect of Photographs of the Handicapped on Donation to Charity: When a Thousand Words May be too Much. Journal of Applied Social Psychology. 1979;9(5):426–31.

35. Evans AT, Peters E, Strasser AA, Emery LF, Sheerin KM, Romer D. Graphic warning labels elicit affective and thoughtful responses from smokers: results of a randomized clinical trial. PloS one. 2015;10(12):e0142879.

36. Brewer NT, Parada Jr H, Hall MG, Boynton MH, Noar SM, Ribisl KM. Understanding why pictorial cigarette pack warnings increase quit attempts. Annals of Behavioral Medicine. 2019;53(3):232–43.

